# COVID-19: Effects of environmental conditions on the propagation of respiratory droplets

**DOI:** 10.1101/2020.05.24.20111963

**Authors:** Lei Zhao, Yuhang Qi, Paolo Luzzatto-Fegiz, Yi Cui, Yangying Zhu

**Author notes:** To whom correspondence should be addressed Yangying Zhu.

## Abstract

As Coronavirus disease 2019 (COVID-19) continues to spread, a detailed understanding on the transmission mechanisms is of paramount importance. The disease transmits mainly through respiratory droplets and aerosol. Although models for the evaporation and trajectory of respiratory droplets have been developed, how the environment impacts the transmission of COVID-19 is still unclear. In this study, we investigate the propagation of respiratory droplets and aerosol particles generated by speech under a wide range of temperature (0 °C to 40 °C) and relative humidity (0% to 92%) conditions. We show that droplets can travel three times farther in low temperature and high humidity environment, while the amount of aerosol increases in high temperature and low humidity environment. The results also underscore the importance of proper ventilation, as droplets and aerosol spread significantly farther in airstreams. This study contributes to the understanding of the environmental impact on COVID-19 transmission.

---

Coronavirus disease 2019 (COVID-19) is an ongoing global pandemic with more than 18 million confirmed cases and over 0.7 million deaths as of August 6th^1^. The disease is caused by the severe acute respiratory syndrome coronavirus 2 (SARS-CoV-2).^2-5^ Among known transmitting pathways of SARS-CoV-2,^6,7^ transmission via respiratory droplets and aerosol is believed to be a primary mode^8,9^. As influenza usually vanishes in the summer^10,11^ and the SARS epidemic was effectively contained in the summer of 2003,^12^ there have been earlier speculations that the current pandemic of COVID-19 may well ebb as the weather warms.^13^ However, the number of confirmed cases of COVID-19 continues to rise rapidly as of August^1^. Therefore, a detailed understanding on the transmission mechanisms of SARS-CoV-2 under different environmental conditions is of paramount importance.

In general, environmental factors can impact the transmission of respiratory diseases through affecting the infectivity of the pathogens^14,15^ and the propagation of respiratory droplets.^16–18^ A hypothesis for the sustained high transmissibility of COVID-19 even in the summer is that SARS-CoV-2 is more persistent at high temperatures compared to influenza and SARS. A number of studies have investigated the resistance of incubated and aerosolized SARS-CoV-2 to heat and humidity.^19-21^ Although more studies are needed to confirm the activity of SARS-CoV-2 in different environments, the virus can generally remain infectious from several minutes to longer than a day in various environments, much longer than the traveling time of respiratory droplets to reach another person through speech or sneeze.^22,23^ Therefore, understanding how environmental conditions impact the propagation of respiratory droplets becomes increasingly important.

The propagation of respiratory droplets plays a critical role in delivering pathogen-carrying agents to susceptible hosts. Respiratory droplets are generated by talking, coughing and sneezing, with initial speeds ranging from ~1 m/s to >100 m/s.^23-25^ Extensive studies have been conducted to investigate the formation,^17,18^ traveling,^16-18,25,26^ and infectivity ^27,28^ of respiratory droplets. The airborne spread of small respiratory droplets and droplet nuclei under different indoor configurations and ventilation designs have been studied to evaluate the indoor infections.^29-31^ Predictions of the infection probability under certain circumstances have been reported as well.^32-34^ These past studies suggest that, during the spread of respiratory droplets, both aerodynamics and evaporation determine the effectiveness of virus propagation. Although models for the evaporation and trajectory of respiratory droplets have been developed,^26,35,36^ how the environmental temperature, humidity and air velocity impact the transmission of COVID-19 is still unclear. Under certain environmental conditions, how far can the virus carriers travel on average? What fraction of droplets will turn into aerosol particles? What role do the HVAC and air conditioning systems play in virus propagation? Quantitative answers to these practical questions can provide urgently needed guidance to both policy makers and the general public, e.g. on environment-specific social distancing rules.

In this study, we investigate the influence of environmental conditions including temperature, humidity and airflow velocity, on the propagation of speech respiratory droplets. We solve the kinematic equations to analyze the transport, accumulation and deposition of respiratory aerosol particles, and calculate the spreading distance of speech droplets based on previous modeling frameworks^26,35,36^ for the evaporation and trajectory of a single droplet. We evaluate the results under a wide range of temperature (0 °C to 40 °C) and relative humidity (0% to 92%) conditions, including weather conditions based on US geographical locations. The results suggest that droplets travel farther in low temperature and high humidity environments, while the amount of aerosol particles increase in high temperature and low humidity environments. In particular, the 6 feet of physical distance recommended by the Centers for Disease Control and Prevention (CDC) is insufficient to eliminate all possible droplet contacts in certain cold and humid environments. The risk of aerosol transmission may be increased in summer, as hot and dry environments facilitate the accumulation of PM2.5. This study highlights the need for adaptive public health precautions based on seasonal weather variations and the local environment, and contributes to the understanding of the environmental impact of COVID-19 transmission.

Figure 1 presents the fate of respiratory droplets. Once released, the droplets begin to evaporate while moving under various forces (gravity, buoyancy and air drag).^26,35,36^ As a result, *large droplets* can directly land on another person, and small droplets dehydrate and become solid *aerosol particles* containing pathogens, salts, enzymes, cells, and surfactants (Figure 1). The *aerosol particles* defined here are different from the airborne aerosol defined by the World Health Organization (WHO),^37^ which employed a straight 5 μm cut-off. Aerosol particles are capable of causing long-range infection because of their long suspension time in air.^21,38,39^ On the other hand, the infection range of *large droplets* is limited to a relatively short distance, because the droplets can directly land on the upper body of another person before drying. Therefore, we define a critical distance L_max_ as the maximum horizontal distance that all respiratory droplets can travel before they either shrink to aerosol particles or descend to the level of another person’s hands (H/2 from the ground, where H is the person’s height, Figure S5).

**Figure 1.**
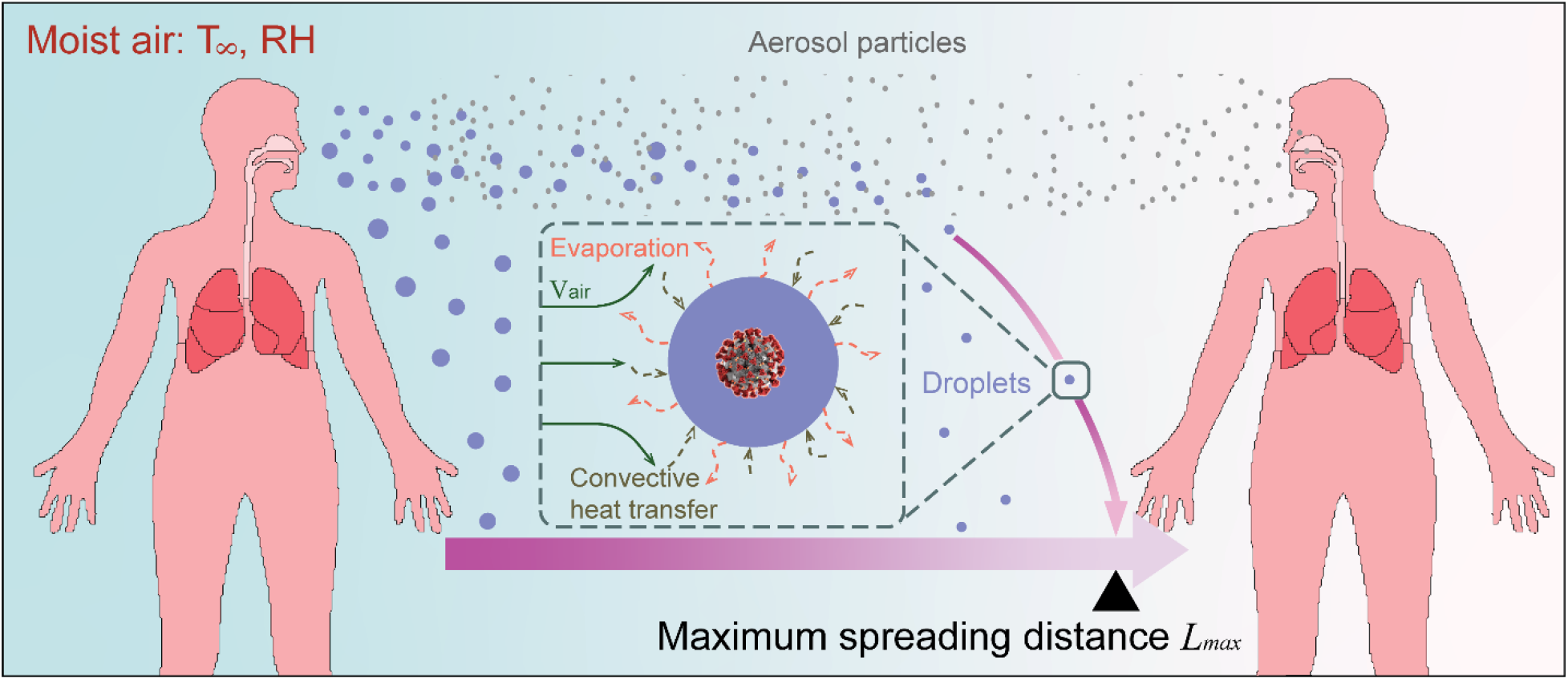
Transmission of COVID-19 through droplets and aerosol particles. After being exhaled by a patient, respiratory droplets with various sizes will travel and simultaneously evaporate in the ambient environment. Small-sized droplets dry immediately to form a cloud of aerosol particles. These particles will suspend in the air for a significant amount of time. Largesized droplets can reach a limited distance and fall to the ground due to gravity. We define L_max_ as the maximum horizontal distance that droplets can travel before they either become dry aerosol particles or descend below the level of another person’s hands, *i.e*., H/2 from the ground, where H is the height of another person. SARS-CoV-2 image by Alissa Eckert, MS; Dan Higgins, MAMS from the Centers for Disease Control and Prevention, USA.

Understanding how the environment affects transmission of COVID-19 requires predicting L_max_ as well as the amount of aerosol particles generated. Before a respiratory droplet evolve into aerosol particle, the evaporation dynamics and the trajectory of the droplet have been modeled by previous works^26,35,36^, based on aerodynamics, kinematics and heat and mass transfer. These modeling frameworks are used in this study to predict L_max_ and aerosolization rate φ_a_ (defined as the percentage of droplets that become aerosol particles) for a cluster of speech droplets in different environments. After respiratory droplets completely dehydrate, we solve the kinematic equations to analyze the transport, accumulation and deposition of residual aerosol particles. The detailed model formulations, including the dynamic analysis on droplets, Brownian motion, and kinematic analysis on aerosol particles, are provided in the Supporting Information.

Key parameters considered in this study include the initial droplet velocity v_0_, the distribution of initial droplet size d_0_, environmental temperature T_∞_, relative humidity RH, and background air velocity V_air_. The initial velocity v_0_ of droplets was taken as 4.1 m/s for speaking.^40^ We focus on modeling speech droplets to mimic a social distancing situation where people keep a reasonable physical distance and only sneeze/cough into a tissue or their elbows. The probability distribution of initial droplet diameter d_0_ is derived from a previous work.^418^ The initial temperature T_0_ of exhaled droplets was set to be 33°C.^429^ For the environment, we used V_air_ = 0.3 m/s in horizontal direction as the wind speed for a typical indoor environment,^43^ and varied V_air_ from 0-3 m/s when analyzing the effect of wind. Respiratory droplets are assumed as a physiological saline solution (0.9% weight fraction)^26^ for simplicity. Even though most buildings in the US are maintained at 21-24 °C all year round,^44^ many regions in the world do not have proper air-conditioning systems. In addition, the temperature and humidity of some industrial settings such as meat processing plants are maintained outside typical setpoints of residential buildings and superspreading events have been reported therein. Therefore, we vary environmental temperature T_„_ and relative humidity RH from 0-42°C and 0-0.92, respectively, to include diverse situations.

The results are presented in Figure 2. For transmission via droplet contact, we plotted L_max_ as defined previously, under different temperature and humidity conditions. Figure 2a shows that droplets can travel a longer distance in humid and cool environments. Therefore, such environments require a longer physical distance. In extremely cold and humid scenarios, droplets can reach as far as 6 m. On the other hand, respiratory droplets evaporate faster in hot and dry environments (Figure S4). As those droplets sharply decrease in size, the horizontal traveling distance is reduced due to the growing damping effect of air. In most regions of Figure 2(a), L_max_ exceeds 1.8 m, *i.e*., the 6 feet of physical distance recommended by CDC. Therefore, current social distancing guidelines may be insufficient in preventing transmission via droplet contact, especially for cold and humid environments.

**Figure 2.**
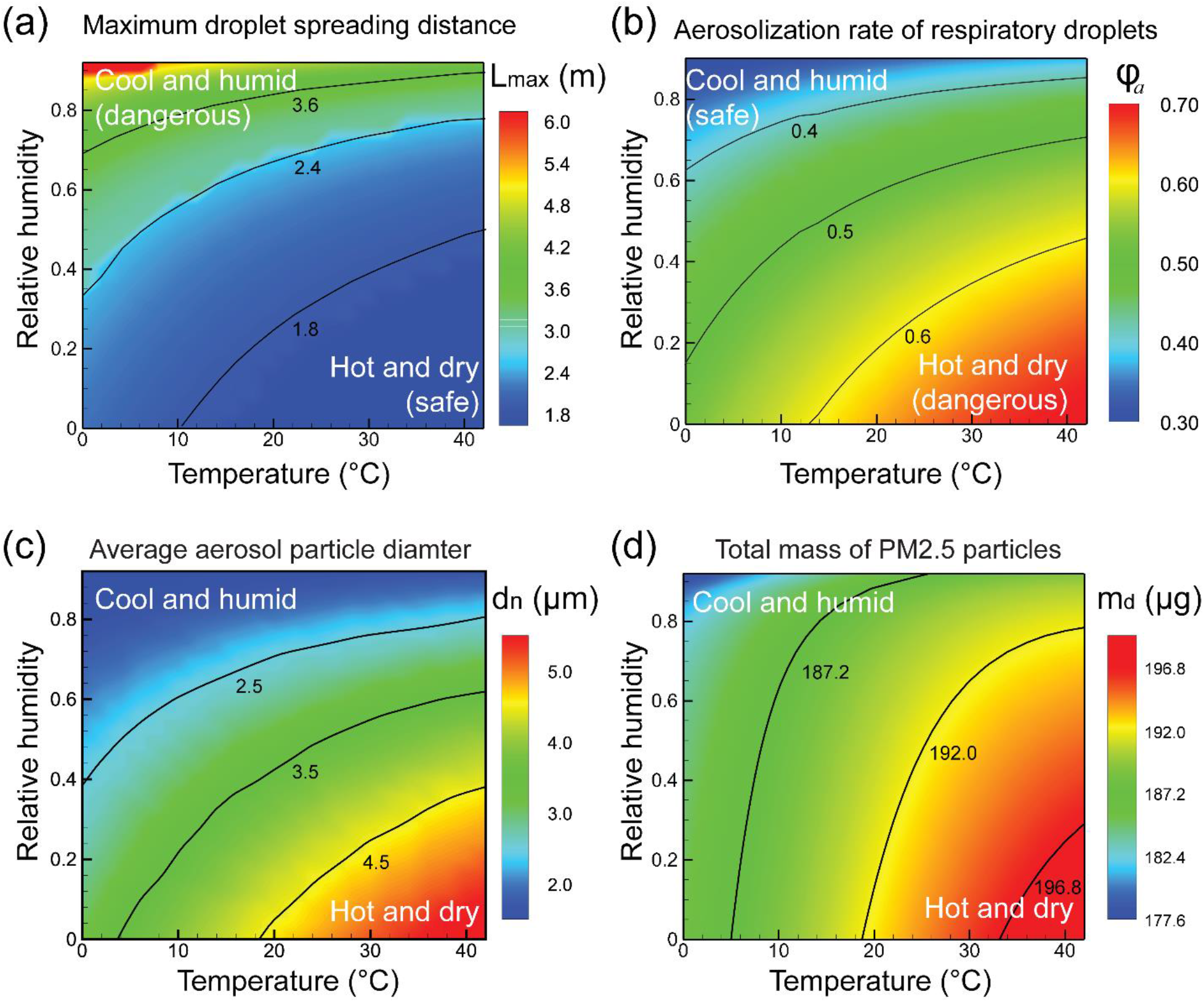
Effect of environmental factors on the transmission of COVID-19 via means of droplets contact and exposure to aerosol particles, respectively. (a) The maximum droplet traveling distance L_max_ under different weather conditions in terms of temperature and relative humidity. Droplets can reach a longer distance in a cool and humid environment. (b) Aerosolization rate φ_a_, defined as the percentage of respiratory droplets turning into aerosol particles that can potentially travel beyond L_max_, under different weather conditions in terms of temperature and humidity. (c) The average diameter of completely dry aerosol particles, under different weather conditions. (d) The total mass of PM2.5 floating in air that are produced by respiratory droplets per person at steady state in an enclosed space.

A quick remark regarding droplet-based transmission is that, if we consider a relaxed standard that only blocks 95% of viruses carried by respiratory droplets (excluding viruses carried by aerosol particles), this distance occurs at 1.4 m based on our results (Figure S7). Here we assumed a constant concentration of the virus in the droplets and the number of viruses is consequently proportional to the initial mass of respiratory droplets.^45^ Interestingly, we find that this relaxed social distancing standard does not show an apparent dependence on the environment (see Supporting Information). Therefore, 1.4 m may serve as a relaxed weather-independent criterion that can prevent the majority of viruses from directly landing on another person. Caution is still required as this relaxed rule does not block transmission via aerosol particles and is evaluated for speaking mode with an air velocity of 0.3 m/s. As discussed in a later section, this distance increases for an increased air velocity.

We also predict the aerosolization rate φ_a_ in different environments to evaluate the potential risk of transmission via aerosol particles. This transmission mode can be highly effective,^46^ as evidenced by the increasing number of superspreading events that occurred at indoor environments. On the contrary to the trend observed for L_max_, Figure 2(b) predicts an increasing aerosolization rate φ_a_ for hot and dry environments. The terminal sizes of these aerosol particles are in the range of 1 μm to 15 μm based on our results (Figure 2(c) and Figure S8). Such small particles can potentially suspend in air for hours ^34,38^ before settling (Figure S8) and tend to accumulate in public areas such as schools, offices, hotels, and hospitals. Therefore, the long-range infection induced by aerosol particles deserves more attention in summer, especially in dry weather.

The effectiveness of aerosol transmission also relies on the ability of aerosol particles to infiltrate respiratory tracts. Generally, small particles are able to infiltrate deeper in the respiratory tract to establish infections, as they can travel with inhaled air current and avoid impaction within the nasal region.^27^ In particular, the filtration efficiency of facial masks is also dependent on the particulate diameter and the proportion between aerosol particles and droplets.^47^ Therefore, understanding the transport and size distribution of aerosol particles can provide key information to the design and fabrication of proper facial covering under different circumstances to ensure optimal protection. We calculate the average diameter of aerosol particles floating in air under different weather conditions. The terminal size of an aerosol particle after dehydration is calculated by approximating to the volume of sodium chloride originally dissolved in the aqueous solution. The volume of viruses, estimated based on reported viral load in clinical samples^48^ and the diameter of the virus^49^, is negligible compared to that of the salt contained in respiratory liquids. Therefore, we only considered the salt when modeling the size of aerosol particles (see Supporting Information). Figure 2(c) demonstrates that aerosol particles have average diameters between 2-5 μm, and the maximum diameters are generally smaller than 10 μm, indicative of their strong ability to penetrate into the human respiratory system. The average particle diameter is increased for hot and dry weather, because more large droplets can completely evaporate due to enhanced evaporation (Figure 2b).

Since PM2.5 has been associated with a higher possibility of reaching the lung,^50^ we then discuss the transport and deposition of aerosol particles classified within PM2.5 in different environment. We first computed the size-dependent suspension time t_s_ from the following equation:

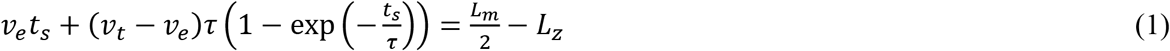

where v_e_ is the falling velocity of the aerosol particle by assuming gravity is entirely balanced by buoyancy and air drag, v_t_ is the downward velocity of the aerosol particle at the time of drying out. If we neglect the Brownian motion, the time constant *τ* of the micro-sized droplet/particle can be written as (see Supporting Information for details on Brownian motion and *τ*):

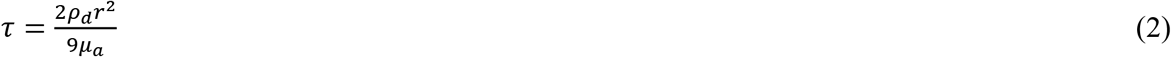

where ρ_d_ is the density of the droplet, r is the droplet radius and μ_a_ is the dynamic viscosity of air. For respiratory droplets with diameters smaller than 100 μm, the time constant *τ* is less than 0.05 s, indicative of a strong damping effect of air. While large aerosol particles can suspend in air for at least 25 minutes, small aerosol suspends substantially longer (Figure S8(b)), which agrees with previous literature.^18^

We further calculate the steady-state total mass m_d_ of PM2.5 produced by one patient that speaks continuously in an enclosed and unventilated space with a volume of V_e_. The generation rate of speech droplets is from a previous study.^51^ The steady state is reached when the number of aerosol particles produced from speaking is equal to the number of particles depositing onto the ground. Given the value of V_e_ and the average number of patients, m_d_ can be used to estimate the number density of pathogen-carrying PM2.5 in air. Figure 2(d) presents m_d_ in different environmental conditions. There is slightly more PM2.5 suspending in air in a hot and dry environment than in a cold and humid environment. This is mainly due to the increased suspension time in hot and dry environments as a result of the corresponding thermophysical properties of the air. By combining Figures 2(b) and 2(d), the hot and dry environment not only increases the percentage of respiratory droplets turning into aerosols, but also facilitates the accumulation of PM2.5 in an enclosed space. These observations raise concerns over aerosol transmission of COVID-19 in summertime, especially when the humidity is low.

Background air velocity is another environmental factor that impact the transmission of contagious diseases. ^52-54^ Here, we investigate the effect of the velocity of a horizontal airflow on the droplet spreading distance and aerosolization rate under a typical environment (*T*_∞_ = 23°C, RH = 0.50). In practice, air velocity distribution can be quite complex, and detailed CFD models are required for a more accurate prediction.^55,56^ For indoor conditions, air velocity is generally maintained below 0.3 m/s for thermal comfort purposes.^57^ We change the air speed from completely stagnant (V_air_ = 0 m/s), to V_air_ = 3 m/s to include diverse environmental settings. For a worst-case estimation, we consider a scenario where the airflow is always directed from a patient to another susceptible host. As shown in Figure 3, the spreading distance of droplets increases dramatically as the airflow velocity increases, which can reach 23 m at V_air_ = 3 m/s. In addition, the spreading distance of aerosol particles is greatly increased as well, since they are smaller in size and travel with the wind. In summary, improper airflow configurations may expand the traveling distance of pathogen-carrying droplets and aerosol particles, although introducing fresh outdoor air can effectively dilute the accumulation of infectious aerosol particles.^52^ This poses stringent requirements on the meticulous design of ventilation configurations in non-hospital facilities, so as to curb the transmission of COVID-19.

**Figure 3.**
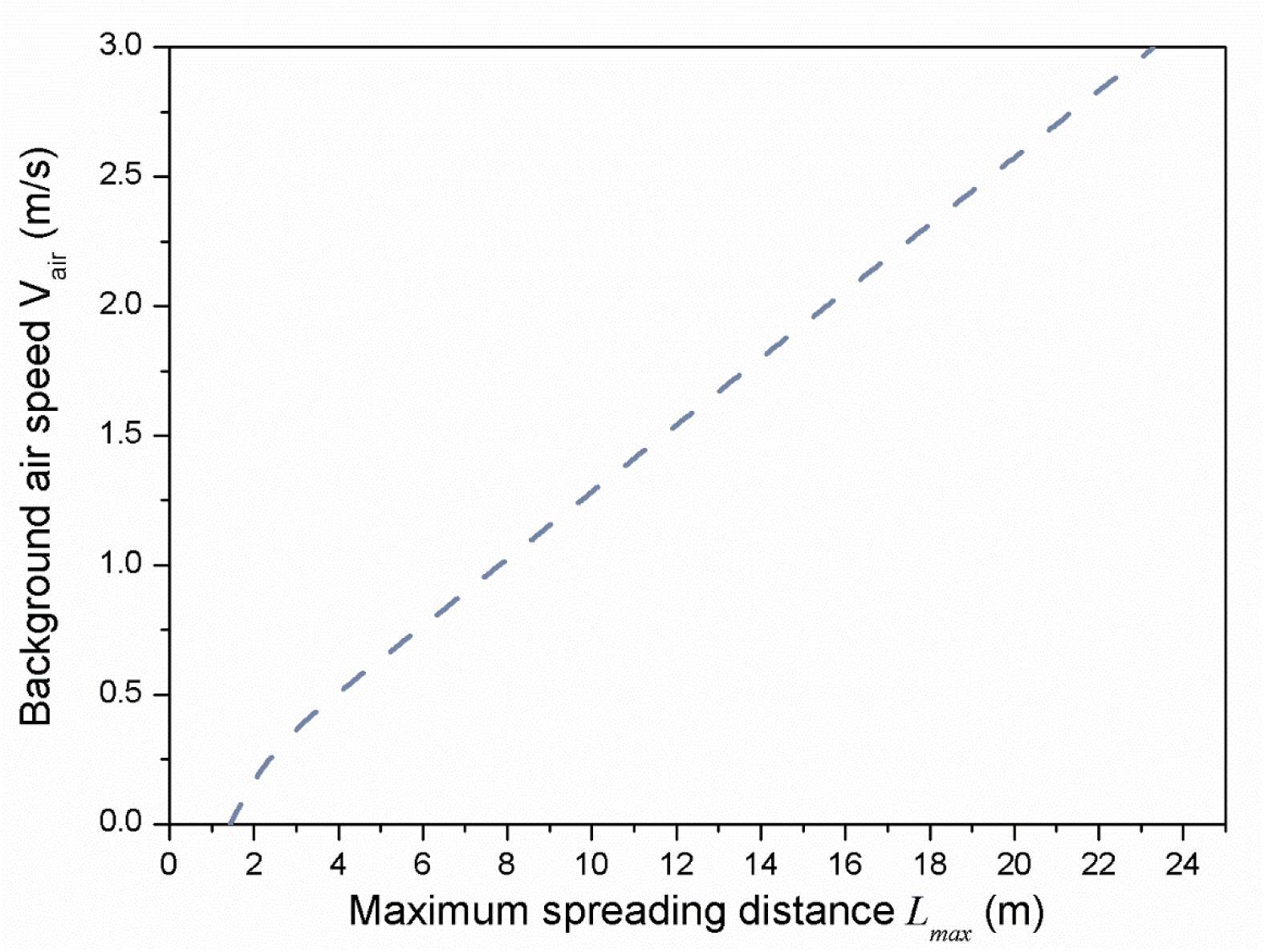
The effect of a horizontal, constant background airflow. Droplet spreading distance *L_max_* as a function of a horizontal, constant air speed.

To help evaluate the risk of infections in different regions across the United States, we compute L_max_ and φ_a_ based on the average weather conditions of each state in August,^58^ and the results are shown Figure 4 (a) and (b). The weather data we collected are a monthly average over day and night as well as rural and urban regions. Figure 4 shows that, the inland regions are more vulnerable to aerosolization of viruses and the coastal regions should be cautious of droplet-based infections. California, on average, exhibits a higher risk of droplet spreading but a lower risk of infection induced by aerosol particles (Figure 4(a) and 4(b)). However, within the state of California, strong variations exist as shown in Figure 4(c) and 4(d). Note that Figure 4(c) and (d) were plotted using the average temperature and relative humidity in the afternoon of August.^58^ We also analyzed L_max_ and φ_a_ in major US cities in both summer (August) and winter (December) climates, as shown in Figure 4(e) and (f). Compared to summer, there is a higher chance of droplet-based infections in winter, while the aerosolization rate merely changes. Considering the extreme stability of SARS-CoV-2 in low temperature,^19^ a recurrent wintertime outbreak of COVID-19 is entirely probable.

**Figure 4.**
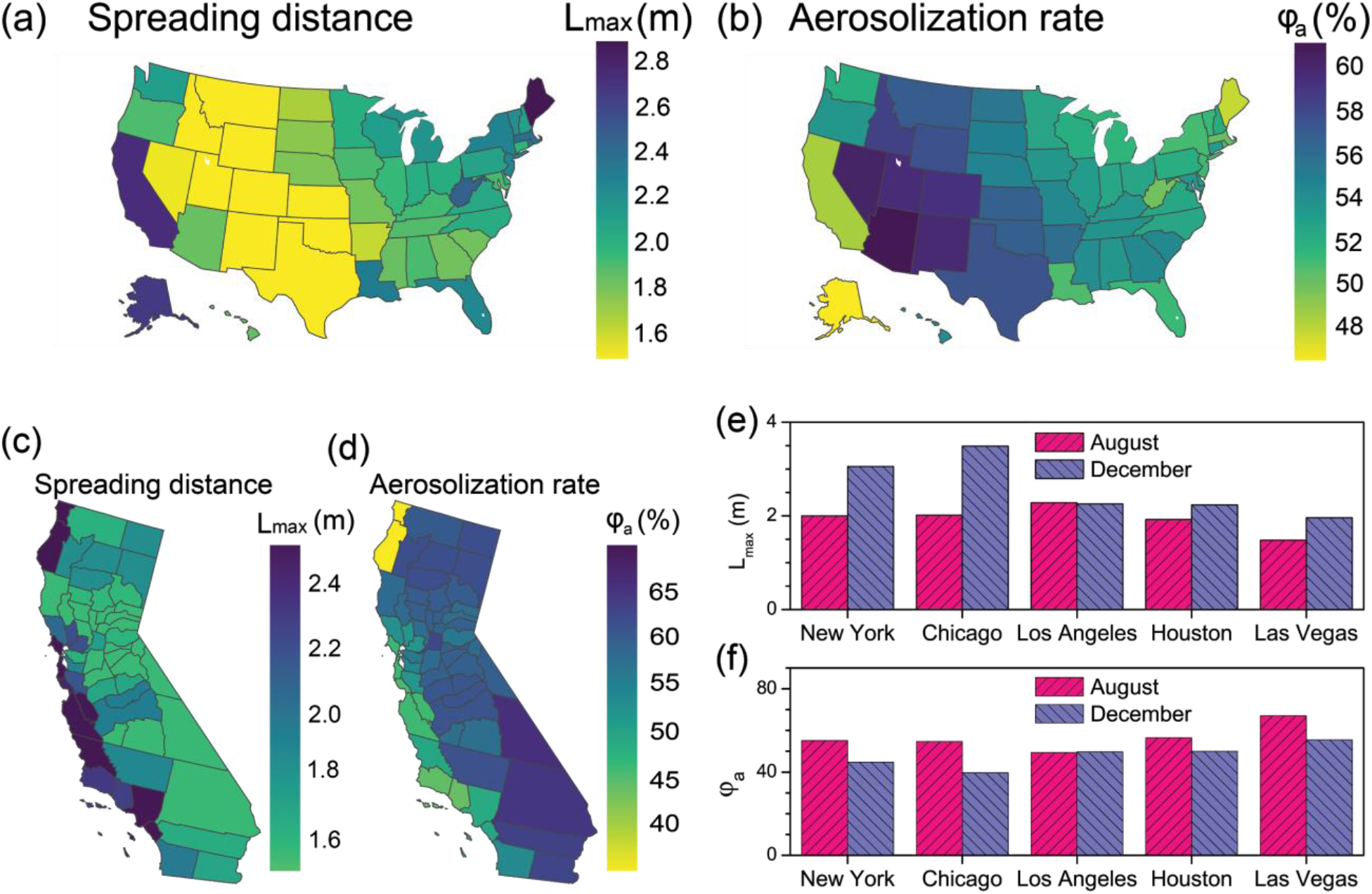
Geographical distribution of droplet spreading distance and aerosolization rate across the United States. (a) Spreading distance L_max_ and (b) aerosolization rate φ_a_ in each US state based on its monthly average weather condition in August. County-by-County distribution of (c) spreading distance L_max_ and (d) aerosolization rate φ_a_ in California based on their monthly average weather condition in August afternoon. (e) Safe distance L_max_ and (f) aerosolization rate φ_a_ in major US cities in summer and winter. Indoor wind speed is assumed (V_air_ = 0.3 m/s).

To summarize, we investigate the effect of environmental temperature, relative humidity and airflow velocity on the propagation of respiratory droplets and accumulation of aerosol particles, in order to elucidate environmental impacts on the transmission of COVID-19. We show that droplets travel farther in low temperature and high humidity environments, while the amount of aerosol particles increase in high temperature and low humidity environments. The current social distancing standard is insufficient in many situations, as droplets may reach as far as 6 m in extreme cold and humid environment. Alternatively, a relaxed standard of 1.4 m is able to block 95% of the viruses carried by respiratory droplets (excluding that carried by aerosol particles) and shows negligible variation over different environmental conditions. Improper airflow can dramatically increase the traveling distance of both droplets and aerosol particles, and consequently increases the risk of COVID-19 transmission. While particulate diameter differs over different environmental settings, proper and adaptive face-covering based on the aerosolization of respiratory droplets is recommended.^59-61^ These findings suggest that adaptive public health measures should be taken in accordance with seasonal weather variations and local environments. The insights gained from this study may shed light on the course of development of the current pandemic, when combined with systematic epidemiological studies.

## Data Availability

The authors confirm that the data supporting the findings of this study are available within the article and its supplementary materials.

## ACKNOWLEDGMENT

This work is supported by the startup funding from University of California, Santa Barbara. We acknowledge synergies made possible by a UCSB VCR COVID-19 Seed Grant to P.L.-F.

## SUPPORTING INFORMATION

The Supporting Information is available free of charge at https://pubs.acs.org Detailed information about the modeling framework, analysis on the Brownian motion, size distribution and evaporation dynamics of respiratory droplets, distance-dependent viral load distribution and calculation of suspending aerosol particles. (PDF)

## Notes

### Competing Interest Statement

The authors have declared no competing interest.

## REFERENCES

(1) Dong, E.; Du, H.; Gardner, L. An Interactive Web-Based Dashboard to Track COVID-19 in Real Time. Lancet Infect. Dis. 2020, 20 (5), 533–534. https://doi.org/10.1016/S1473-3099(20)30120-1

(2) Zhou, P.; Yang, X.-L.; Wang, X.-G.; Hu, B.; Zhang, L.; Zhang, W.; Si, H.-R.; Zhu, Y.; Li, B.; Huang, C.-L.; others. A Pneumonia Outbreak Associated with a New Coronavirus of Probable Bat Origin. Nature 2020, 579 (7798), 270–273.

(3) Wu, F.; Zhao, S.; Yu, B.; Chen, Y.-M.; Wang, W.; Song, Z.-G.; Hu, Y.; Tao, Z.-W.; Tian, J.-H.; Pei, Y.-Y.; others. A New Coronavirus Associated with Human Respiratory Disease in China. Nature 2020, 579 (7798), 265–269.

(4) Guan, W.; Ni, Z.; Hu, Y.; Liang, W.; Ou, C.; He, J.; Liu, L.; Shan, H.; Lei, C.; Hui, D. S.; others. Clinical Characteristics of Coronavirus Disease 2019 in China. N. Engl. J. Med. 2020, 382 (18), 1708–1720.

(5) Letko, M.; Marzi, A.; Munster, V. Functional Assessment of Cell Entry and Receptor Usage for SARS-CoV-2 and Other Lineage B Betacoronaviruses. Nat. Microbiol. 2020, 5 (4), 562–569.

(6) Leung, K.; Wu, J. T.; Liu, D.; Leung, G. M. First-Wave COVID-19 Transmissibility and Severity in China Outside Hubei after Control Measures, and Second-Wave Scenario Planning: A Modelling Impact Assessment. The Lancet 2020, 395(10233), 1382–1393

(7) Arons, M. M.; Hatfield, K. M.; Reddy, S. C.; etc. Presymptomatic SARS-CoV-2 Infections and Transmission in a Skilled Nursing Facility. N. Engl. J. Med. 2020, 382, 2081-2090

(8) Booth, T. F.; Kournikakis, B.; Bastien, N.; etc. Detection of Airborne Severe Acute Respiratory Syndrome (SARS) Coronavirus and Environmental Contamination in SARS Outbreak Units. J. Infect. Dis. 2005, 191 (9), 1472–1477.

(9) Herfst, S.; Schrauwen, E. J.; Linster, M.; etc. Airborne Transmission of Influenza A/H5N1 Virus between Ferrets. Science 2012, 336 (6088), 1534–1541.

(10) Ye, C.; Zhu, W.; Yu, J.; etc. Understanding the complex seasonality of seasonal influenza A and B virus transmission: Evidence from six years of surveillance data in Shanghai, China. Int. J. Infect. Dis. 2019, 81, 57-65.

(11) Lofgren, E.; Fefferman, N. H.; Naumov, Y. N.; etc. Influenza seasonality: underlying causes and modeling theories. J. Virol. 2007, 81(11), 5429–5436.(12)

(12) Likhacheva, A. SARS Revisited. AMA Journal of Ethics 2006, 8(4), 219–222.

(13) Marc Lipsitch, DPhil. Seasonality of SARS-CoV-2: Will COVID-19 go away on its own in warmer weather? https://ccdd.hsph.harvard.edu/will-covid-19-go-away-on-its-own-in-warmer-weather/ (Accessed on May 3rd, 2020).

(14) Lowen, A. C.; Steel, J. Roles of humidity and temperature in shaping influenza seasonality. J. Virol. 2014, 88(14), 7692–7695.

(15) Chan, K. H.; Peiris, J. M.; Lam, S. Y.; etc. The effects of temperature and relative humidity on the viability of the SARS coronavirus. Adv. Virol. 2011, 2011(734690), 1–7.

(16) Bourouiba, L. Turbulent Gas Clouds and Respiratory Pathogen Emissions: Potential Implications for Reducing Transmission of COVID-19. JAMA 2020, 323 (18), 1837–1838.

(17) Mittal, R.; Ni, R.; Seo, J.-H. The Flow Physics of COVID-19. J. Fluid Mech. 2020, 894. https://doi.org/10.1017/jfm.2020.330

(18) Scharfman, B. E.; Techet, A. H.; Bush, J. W. M.; Bourouiba, L. Visualization of Sneeze Ejecta: Steps of Fluid Fragmentation Leading to Respiratory Droplets. Exp. Fluids 2016, 57 (2), 24.

(19) Chin, A.; Chu, J.; Perera, M.; Hui, K.; Yen, H.-L.; Chan, M.; Peiris, M.; Poon, L. Stability of SARS-CoV-2 in Different Environmental Conditions. Lancet Microbe 2020, 1 (1), e10.

(20) van Doremalen, N.; Bushmaker, T.; Morris, D. H.; Holbrook, M. G.; Gamble, A.; Williamson, B. N.; Tamin, A.; Harcourt, J. L.; Thornburg, N. J.; Gerber, S. I. Aerosol and Surface Stability of SARS-CoV-2 as Compared with SARS-CoV-1. N. Engl. J. Med. 2020, 382 (16), 1564–1567.

(21) Fears, A. C.; Klimstra, W. B.; Duprex, P.; Hartman, A.; etc. Comparative Dynamic Aerosol Efficiencies of Three Emergent Coronaviruses and the Unusual Persistence of SARS-CoV-2 in Aerosol Suspensions. 2020, medRxiv 2020.04.13.20063784. medRxiv. https://www.medrxiv.org/content/10.1101/2020.04.13.20063784v1 (Accessed on May 4th, 2020).

(22) Stadnytskyi, V.; Bax, C. E.; Bax, A.; Anfinrud, P. The Airborne Lifetime of Small Speech Droplets and Their Potential Importance in SARS-CoV-2 Transmission. Proc. Natl. Acad. Sci. 2020, 117(22), 11875–11877.

(23) Xie, X.; Li, Y.; Sun, H.; Liu, L. Exhaled Droplets Due to Talking and Coughing. J. R. Soc. Interface 2009, 6, S703-S714.

(24) Cole, E. C.; Cook, C. E. Characterization of Infectious Aerosols in Health Care Facilities: An Aid to Effective Engineering Controls and Preventive Strategies. Am. J. Infect. Control 1998, 26 (4), 453–464.

(25) Chao, C. Y. H.; Wan, M. P.; Morawska, L.; Johnson, G. R.; Ristovski, Z. D.; Hargreaves, M.; Mengersen, K.; Corbett, S.; Li, Y.; Xie, X.; Katoshevski, D. Characterization of expiration air jets and droplet size distributions immediately at the mouth opening. J. Aerosol Sci. 2009, 40(2), 122–133.

(26) Xie, X.; Li, Y.; Chwang, A.; Ho, P.; Seto, W. How Far Droplets Can Move in Indoor Environments–Revisiting the Wells Evaporation-Falling Curve. Indoor Air 2007, 17 (3), 211–225.

(27) Gralton, J.; Tovey, E.; McLaws, M.-L.; Rawlinson, W. D. The Role of Particle Size in Aerosolised Pathogen Transmission: A Review. J. Infect. 2011, 62 (1), 1–13.

(28) Kormuth, K. A.; Lin, K.; Prussin, A. J.; Vejerano, E. P.; Tiwari, A. J.; Cox, S. S.; Myerburg, M. M.; Lakdawala, S. S.; Marr, L. C. Influenza Virus Infectivity Is Retained in Aerosols and Droplets Independent of Relative Humidity. J. Infect. Dis. 2018, 218 (5), 739–747.

(29) Chen, C.; Zhao, B. Some questions on dispersion of human exhaled droplets in ventilation room: answers from numerical investigation. Indoor Air 2010, 20(2), 95–111.

(30) Mui, K. W.; Wong, L. T.; Wu, C. L.; Lai, A. C. Numerical modeling of exhaled droplet nuclei dispersion and mixing in indoor environments. J. Hazard. Mater. 2009, 167(1-3), 736-744.

(31) Ai, Z. T.; Melikov, A. K. Airborne spread of expiratory droplet nuclei between the occupants of indoor environments: A review. Indoor Air 2018, 28(4), 500–524.

(32) Nicas, M.; Best, D. A Study Quantifying the Hand-to-Face Contact Rate and Its Potential Application to Predicting Respiratory Tract Infection. J. Occup. Environ. Hyg. 2008, 5 (6), 347–352.

(33) Nicas, M.; Sun, G. An Integrated Model of Infection Risk in a Health-Care Environment. Risk Anal. 2006, 26 (4), 1085–1096.

(34) Tellier, R. Aerosol Transmission of Influenza A Virus: A Review of New Studies. J. R. Soc. Interface 2009, 6, S783-S790.

(35) Wells, W. On Air-Borne Infection: Study II. Droplets and Droplet Nuclei. Am. J. Epidemiol. 1934, 20 (3), 611–618.

(36) Kukkonen, J.; Vesala, T.; Kulmala, M. The Interdependence of Evaporation and Settling for Airborne Freely Falling Droplets. J. Aerosol Sci. 1989, 20 (7), 749–763.

(37) World Health Organization. Infection Prevention and Control of Epidemic-and Pandemic-Prone Acute Respiratory Infections in Health Care. https://www.who.int/csr/bioriskreduction/infection_control/publication/en/ (Accessed on May 5th, 2020)

(38) Alsved, M.; Bourouiba, L.; Duchaine, C.; Löndahl, J.; Marr, L. C.; Parker, S. T.; Prussin, A. J.; Thomas, R. J. Natural Sources and Experimental Generation of Bioaerosols: Challenges and Perspectives. Aerosol Sci. Technol. 2020, 54 (5), 547–571.

(39) Poulain, S.; Bourouiba, L. Disease Transmission via Drops and Bubbles. Phys. Today 2019, 72 (5), 70–71.

(40) Kwon, S.-B.; Park, J.; Jang, J.; Cho, Y.; Park, D.-S.; Kim, C.; Bae, G.-N.; Jang, A. Study on the Initial Velocity Distribution of Exhaled Air from Coughing and Speaking. Chemosphere 2012, 87 (11), 1260–1264.

(41) Duguid, J. P. The Size and the Duration of Air-Carriage of Respiratory Droplets and Droplet-Nuclei. Epidemiol. Infect. 1946, 44 (6), 471–479.

(42) Höppe, P. Temperatures of Expired Air under Varying Climatic Conditions. Int. J. Biometeorol. 1981, 25 (2), 127–132.

(43) Baldwin, P. E.; Maynard, A. D. A Survey of Wind Speeds in Indoor Workplaces. Ann. Occup. Hyg. 1998, 42 (5), 303–313.

(44) Hoyt, T., Arens E., and Zhang H. Extending air temperature setpoints: Simulated energy savings and design considerations for new and retrofit buildings. Build. Sci. 1998, 88, 8996.

(45) Vejerano, E. P.; Marr, L. C. Physico-Chemical Characteristics of Evaporating Respiratory Fluid Droplets. J. R. Soc. Interface 2018, 15 (139), 20170939.

(46) Bahl, P.; Doolan, C.; de Silva, C.; Chughtai, A. A.; Bourouiba, L.; MacIntyre, C. R. Airborne or Droplet Precautions for Health Workers Treating COVID-19? J. Infect. Dis. 2020, https://doi.org/10.1093/infdis/jiaa189

(47) Shakya, K. M.; Noyes, A., Kallin, R.; Peltier, R. E. Evaluating the efficacy of cloth facemasks in reducing particulate matter exposure. J. Exposure Sci. Environ. Epidemiol. 2017, 27(3), 352–357.

(48) Wölfel, R.; Victor, M. C.; Wolfgang, G.; Michael, S.; Sabine, Z.; Marcel, A. M.; Daniela, N. et al. Virological assessment of hospitalized patients with C0VID-2019. Nature 2020, 581, 465-469.

(49) Bar-On, Y. M.; Flamholz, A.; Phillips, R.; Milo, R. Science Forum: SARS-CoV-2 (COVID-19) by the numbers. Elife 2020, 9, e57309.

(50) Xing, Y.-F.; Xu, Y.-H.; Shi, M.-H.; Lian, Y.-X. The Impact of PM2. 5 on the Human Respiratory System. J. Thorac. Dis. 2016, 8 (1), E69.

(51) Asadi, S.; Wexler, A. S.; Cappa, C. D.; Barreda, S.; Bouvier, N. M.; Ristenpart, W. D. Aerosol emission and superemission during human speech increase with voice loudness. Sci. Rep. 2019, 9(1), 1–10.

(52) Cedeño-Laurent, J. G.; Williams, A.; MacNaughton, P.; Cao, X.; Eitland, E.; Spengler, J.; Allen, J. Building Evidence for Health: Green Buildings, Current Science, and Future Challenges. Annu. Rev. Public Health 2018, 39 (1), 291–308.

(53) Li, Y.; Leung, G. M.; Tang, J. W.; Yang, X.; Chao, C. Y.; Lin, J. Z.; Lu, J. W.; Nielsen, P. V.; Niu, J.; Qian, H. Role of Ventilation in Airborne Transmission of Infectious Agents in the Built Environment-a Multidisciplinary Systematic Review. Indoor Air 2007, 17 (1), 2–18.

(54) Ai, Z. T.; Huang, T.; Melikov, A. K. Airborne Transmission of Exhaled Droplet Nuclei between Occupants in a Room with Horizontal Air Distribution. Build. Environ. 2019, 163, 106328.

(55) Li, Y.; Nielsen, P. V. CFD and ventilation research. Indoor Air 2011, 21(6), 442–453.

(56) Ramponi, R., Blocken, B. CFD simulation of cross-ventilation for a generic isolated building: impact of computational parameters. Build. Sci. 2012, 53, 34-48.

(57) Owen, M. S. ASHRAE Handbook: HVAC systems and equipment. American Society of Heating, Refrigerating and Air-Conditioning Engineers, Inc., Atlanta, 2004.

(58) Weather Averages for the United States. https://www.currentresults.com/Weather/US/weather-averages-index.php (Accessed on May 7th, 2020)

(59) Liao, L.; Xiao, W.; Zhao, M.; Yu, X.; Wang, H.; Wang, Q.; Chu, S.; Cui, Y. Can N95 Respirators Be Reused after Disinfection? How Many Times? ACS Nano 2020, 14(5), 6348–6356

(60) Zhao, M.; Liao, L.; Xiao, W.; Yu, X.; Wang, H.; Wang, Q.; Lin, Y. L.; Kilinc-Balci, F. S.; Price, A.; Chu, L.; Chu, M. C.; Chu S.; Cui, Y. Household materials selection for homemade cloth face coverings and their filtration efficiency enhancement with triboelectric charging. Nano Letters 2020, 20(7), 5544–5552.

(61) Konda, A.; Prakash, A.; Moss, G. A.; Schmoldt, M.; Grant, G. D.; Guha, S. Aerosol filtration efficiency of common fabrics used in respiratory cloth masks. ACS Nano 2020, 14(5), 63396347

